# Integration of rare large-effect expression variants improves polygenic risk prediction

**DOI:** 10.1101/2020.12.02.20242990

**Authors:** Craig Smail, Nicole M. Ferraro, Matthew G. Durrant, Abhiram S. Rao, Matthew Aguirre, Xin Li, Michael J. Gloudemans, Themistocles L. Assimes, Charles Kooperberg, Alexander P. Reiner, Qin Hui, Jie Huang, Christopher J. O’Donnell, Yan V. Sun, Million Veteran Program, Manuel A. Rivas, Stephen B. Montgomery

**Author notes:** Corresponding authors: Craig Smail, Stephen Montgomery (lead contact).

## Abstract

Polygenic risk scores (PRS) aim to quantify the contribution of multiple genetic loci to an individual’s likelihood of a complex trait or disease. However, existing PRS estimate genetic liability using common genetic variants, excluding the impact of rare variants. We identified rare, large-effect variants in individuals with outlier gene expression from the GTEx project and then assessed their impact on PRS predictions in the UK Biobank (UKB). We observed large deviations from the PRS-predicted phenotypes for carriers of multiple outlier rare variants; for example, individuals classified as “low-risk” but in the top 1% of outlier rare variant burden had a 6-fold higher rate of severe obesity. We replicated these findings using data from the NHLBI Trans-Omics for Precision Medicine (TOPMed) biobank and the Million Veteran Program, and demonstrated that PRS across multiple traits will significantly benefit from the inclusion of rare genetic variants.

## Introduction

A major goal of complex disease genetics is predicting an individual’s disease risk. Recent efforts have aimed at summarizing genome-wide risk for multiple traits and diseases using polygenic risk scores (PRS)^1–6^, which are derived by summing genome-wide common genetic variants associated with a given phenotype. PRS have demonstrated stratification of genetic disease risk, but there remains substantial unexplained variability in these predictions. One potential explanation for this variability is the presence of rare variants with large phenotypic effects that are unaccounted for in PRS models^2^.

However, despite known contributions of rare genetic variants to complex traits and diseases^7,8^, rare variants are difficult to robustly characterize and integrate into PRS predictions due to their abundance in the genome, poor interpretability and sample size constraints. To in-part alleviate this challenge, it has previously been shown that individuals with outlier gene expression have an increased burden of rare variants proximal to the outlier gene^9–12^, and that this subset of rare variants tend to have larger effects on traits and diseases^13,14^.

Given the known large effects of rare variants linked to expression outliers - and that these variants are not currently included in existing PRS - we sought to test whether this subset of rare variants can aid in explaining instances where an individual’s phenotype deviates from their phenotype as predicted by their PRS. We present an approach that summarizes the phenotypic effects in UKB^15^ of an increasing burden of rare variants associated with outlier gene expression discovered in GTEx. We focus primarily on body mass index (BMI) and obesity given the growing public health emergency of severe obesity in the US and around the world^16^, the availability of high-quality publicly-available PRS for BMI, known polygenicity, and sample size considerations.

## Results

### Identification of rare, large-effect expression variants

To identify rare variants linked to gene expression outliers that could also be tested for their effects on complex traits, we intersected the set of single nucleotide variants with gnomAD^17^ minor allele frequency (MAF) > 0 and < 1% identified in GTEx v7 with high-quality imputed variants in the UKB (**Fig. 1A**). From a starting set of 6,134,805 unique rare variants, we identified 1,307,023 (21.3%) variants also within the UKB (**Fig. 1B**). From this intersecting set, we compared the set of variants found in GTEx outlier to non-outlier individuals to isolate the subset of rare variants present in gene expression outlier individuals only. This process was conducted in two ways; “top-outlier” where only rare variants from the most extreme outlier individual(s) (maximum of two individuals per gene - most under-expressed and most over-expressed individuals, abs(Z)>2), and “all outliers” where all rare variants from individuals with abs(Z)>2 were included (**Methods; Sup. Table 1**). Rare variants found in both outlier and non-outlier individuals were subsequently removed. Variants were then linked to a specific gene if they fell within the gene body or +/- 10 Kb (N genes: “top-outlier” = 3,732; “all-outliers” = 15,095). We consider the “top-outlier” as a high-confidence set, given that these variants are found in the most extreme expression outliers. The “all-outlier” method allows us to expand the range of variants, guided by the properties of the “top-outlier” variant set. We further defined a corresponding set of non-outlier/control variants, matched on both the gnomAD MAF and CADD^18^ scores of outlier variants (**Methods**).

**Figure 1.**
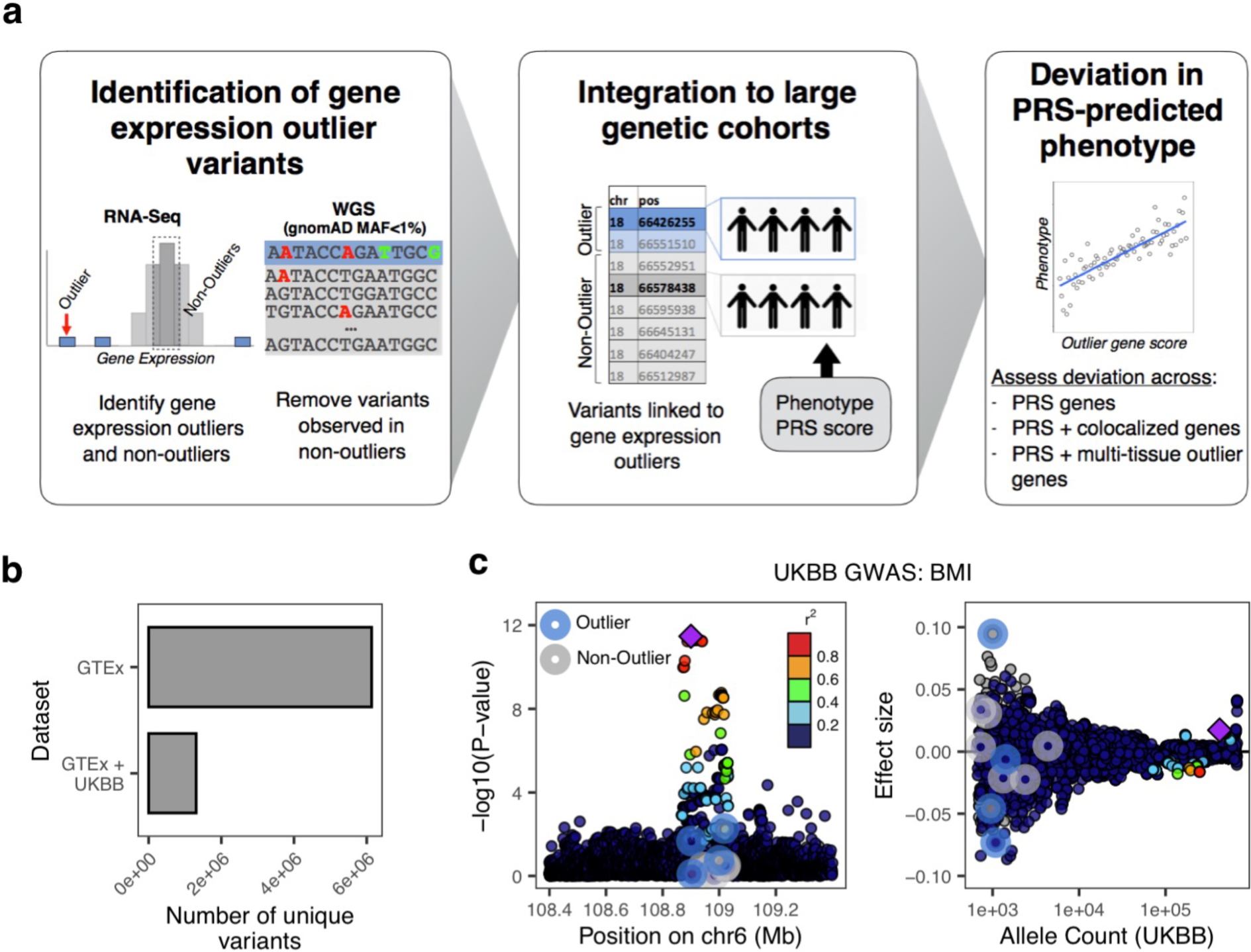
Linking rare variants associated with gene expression outliers in GTEx to large-scale genetic cohorts. **A**. Overview of methodology: rare variants (gnomAD MAF<1%) were identified in expression outlier samples across 48 GTEx tissues, discarding any variants also observed in non-outliers; variants from non-outlier samples were selected per gene, matched on gnomAD MAF (<1%) and CADD score (+/- 5), to create a control set; variants were linked to UKB samples, combined with UKB samples’ BMI PRS, ancestry PCs, age, sex, and phenotype data. **B**. Number of rare GTEx variants recovered in UKB imputed set. **C**. Example gene locus (*FOXO3*) containing a common variant genome-wide significant hit for BMI illustrates large GWAS effect size of outlier-associated variants: (left) showing distribution of - log10(P-values) for UKB BMI GWAS for all outlier (blue halo) and non-outlier (gray halo) variants linked to the gene; (right) associated effect sizes, stratified by UKB allele count. Outlier variants have among the largest effects in gene locus but do not reach genome-wide significance. Points are colored by LD (1000 Genomes phase 3, European samples) relative to lead variant (top P-value) (purple diamond) in gene locus.

We observed that individuals were often carriers for multiple outlier rare variants. Considering a sample cohort of individuals from UKB (N = 120,944) (**Methods; Sup. Fig. 1**), each individual had an average of 23 (“top-outlier”) and 304 (“all-outlier”) outlier variants. To evaluate if these variants cumulatively were biased in effect direction (i.e. risk or protective) for a highly polygenic trait, we assessed UKB BMI GWAS effect directions and observed no significant differences. On average, individuals carried 11 potential protective rare variants and 12 potential risk variants using the “top-outlier” approach.

### Large-effect, rare expression variants impact BMI and obesity in UK Biobank

To evaluate if rare expression outlier-associated variants had greater effect sizes than matched non-outlier variants (used as a control set), we focused on BMI and obesity GWAS from the UKB. We observed that a subset of outlier variants had relatively larger GWAS effects; for example, an outlier variant linked to the gene *FOXO3* has an effect size rank of 1/3059 in a 1 Mb locus (centered on the top genome-wide significant variant) (**Fig. 1C**), and among the top 0.07% of effect sizes overall, across all variants measured across the UKB for BMI.

To systematically assess whether these outlier variants had higher effect sizes than non-outlier variants, we performed a permutation test (N permutations = 10,000) using outlier (“top-outlier”; n variants = 8,272) and matched non-outlier variants (n variants = 29,659) that fall within 10kb of any PRS variant to assess how often randomly-drawn outlier variants had larger effect sizes than non-outlier variants. For BMI GWAS, we observed a mean odds ratio of 1.02 when comparing outlier vs. non-outlier variants, and a mean odds ratio of 1 when comparing non-outlier variants to themselves (Wilcoxon test, *P* < 1×10^−16^). For obesity GWAS (ICD-10 E66), we observed an increased mean odds ratio of 1.1 (Wilcoxon test, *P* < 1×10^−16^) (**Fig. 2A**). When increasing the outlier expression Z-score threshold, we observed progressively larger odds ratios (mean odds ratio (BMI GWAS): abs(Z)>4 = 1.28; abs(Z)>6 = 1.58), but not when comparing non-outlier variants only (mean odds ratio (BMI GWAS): abs(Z)>4 = 1; abs(Z)>6 = 1) (Wilcoxon test, *P* < 1×10^−16^ for both comparisons) (**Fig. 2B**). We replicated the permutation test findings using a subset of the same rare variants that are also available in the Million Veteran Program (MVP) BMI GWAS (N variants: outlier = 4,955; non-outlier = 18,145), and observed similar results (mean odds ratio: outlier vs. non-outlier = 1.05; non-outlier only = 1; *P* < 1×10^−16^) (**Fig. 2C**). We further directly compared effect sizes between outlier and control variants and observed significantly increased effect sizes for outlier variants that increased with outlier Z-score thresholds (**Fig. 2D**; Ansari test, *P* < 1×10^−16^ for all comparisons).

**Figure 2.**
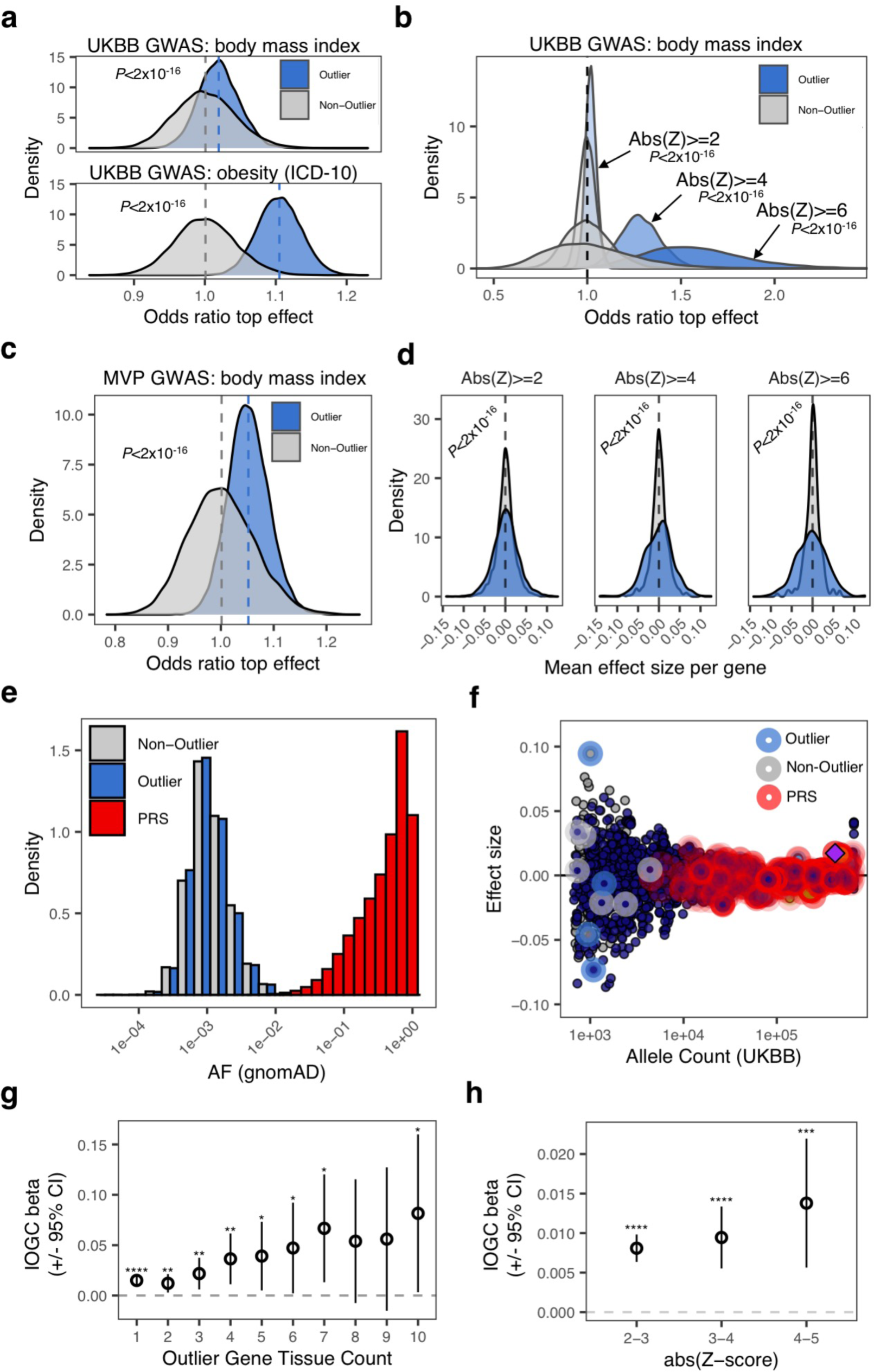
Characterizing outlier and non-outlier variants using large-scale GWAS. **A**. Distribution of odds ratios from permutation testing (N permutations = 10,000) to assess relative effect size comparing outlier- and non-outlier variants per gene in UKB GWAS for BMI (top) and obesity (bottom). Across each permutation, the absolute effect size for a randomly-chosen outlier sample and matched non-outlier sample was obtained for each gene and summed in a contingency matrix to quantify the number of genes where the outlier variant had an absolute effect size greater than the non-outlier variant (blue shading). This process was repeated for randomly selected non-outlier variants only (gray). P-values were obtained using a Wilcoxon rank sum test. Subset to genes linked to PRS variants. **B**. Distribution of odds ratios from permutation testing (N permutations = 10,000) (permutation testing method as detailed in (A.)), across progressively more-stringent GTEx outlier Z-scores. Odds ratio increases as a function of outlier Z-score. **C**. Distribution of odds ratios from permutation testing (N permutations = 10,000) (permutation testing method as detailed in (A.)), using the Million Veteran Program (MVP) GWAS for BMI. **D**. Dispersion of mean effect sizes per gene for outlier (blue) and non-outlier variants (gray) across genes with variants overlapping a publicly-available PRS for BMI, stratified by GTEx outlier Z-score. P-values were obtained using an Ansari Test. **E**. Distribution of gnomAD allele frequency for outlier-associated (blue), and non-outlier (grey) variants, and variants included in a publicly-available PRS for body mass index (red). Outlier- and non-outlier-associated variants are rarer than variants included in the PRS. **F**. Example gene locus (*FOXO3*) containing a genome-wide significant hit in UKB BMI GWAS (GWAS ID 21001) (purple diamond). Effect sizes for each variant in locus are displayed on the y-axis and UKB allele count for each variant is displayed on x-axis. Points are colored by LD (1000 Genomes phase 3, European cohort). Outlier-associated variants are highlighted in blue, non-outlier-associated variants are highlighted in gray, PRS variants are highlighted in red. Outlier-associated variants have largest effect sizes in locus. PRS variants tend to be common with small effect size. **G**. Coefficient estimate for IOGC score increases when subsetting to outlier-associated variants where variants are identified in outliers in an increasing number of GTEx tissues. X-axis indicates tissue threshold (i.e. tissue count>=N). **H**. Mean change in BMI per unit change in IOGC score at difference Z-score cutoffs. Variants identified in more-severe outliers (by abs(Z-score)) have larger effects on BMI.

We next assessed whether outlier variant effects were concordant with predictions of effect direction from common variant associations. We compared GWAS effect direction between cis-eQTLs and outlier variants for the same loci matched on slope (as an example, positive cis-eQTL slope and over-expression outliers both leading to increased GWAS risk) (**Methods**). We stratified results by cis-eQTL variant GWAS p-value and outlier-associated variant Z-score, observing that variants identified in more-severe (by Z-score) expression outliers have overall better concordance in GWAS effect direction with cis-eQTL variants, across genes. For example, at a cis-eQTL variant GWAS P-value cutoff <= 1×10^−6^, we observed 50, 96, and 100% concordance for variants passing absolute Z-score thresholds of 2, 3, and 4, respectively (**Sup. Fig. 2**).

### Independent outlier gene count (IOGC) score stratifies BMI

To identify the impact of multiple outlier variants in an existing high-quality, publicly-available BMI PRS, we used data from Khera et al. (2019)^1^. We first obtained gnomAD AF for PRS variants, and observed that these PRS alleles have a mean gnomAD AF = 0.49 (SD = 0.29) (**Fig. 2E**). Plotting GWAS effect sizes by UKB allele count for an example locus (gene *FOXO3*) further illustrates that PRS variants tend to be common variants with small effects (**Fig. 2F**). We calculated PRS for each individual in our UKB validation cohort (N = 120,944) and observed the expected gradients in mean BMI and weight increasing by PRS deciles (**Sup. Fig. 3**). We then used a linear regression model to assess change in BMI given an individual’s PRS, sex, age, first ten components of genetic ancestry, genotyping array, and a score that quantifies the total outlier-variant burden per individual, computed by subtracting total protective from total risk outlier-variants collapsed to gene-level (**Methods; Sup. Fig. 4**). We refer to this score as the independent outlier gene count (IOGC) score henceforth. We observed significant coefficient estimates for 10/15 features in the model, including IOGC score (linear regression *r* = 0.015, *P* = 7×10^−7^) (**Sup. Fig. 5**). We computed the rate of concordance in GWAS effect (i.e. risk/protective) and outlier direction for outlier variants linked genes across UKB individuals, observing a rate of concordance of 86.7% for individuals with >=2 outlier variants linked to each gene. This rate of concordance is remarkably stable when increasing the outlier variant threshold per gene (82.2%, 81.8%, 83.2%, for >= 3, 4, and 5 outlier variants, respectively).

We sought to clarify whether IOGC variants identified from outlier individuals in GTEx were driving downstream effects on BMI in excess of what is observed by selecting random subsets of non-outlier rare variants. We investigated this by first selecting random subsets of the matched non-outlier variants (matching the number of outlier variants across permutations, N variants = 8,272; N permutations = 10,000), observing that the IOGC beta estimate when using outlier variants exceeds that which would be expected based on random subsets of non-outlier variants (mean IOGC non-outlier = 0.0075; empirical *P* = 0.0012) (**Sup. Fig. 6A**), validating the findings of the permutation test described in the previous section. Furthermore, when outliers where observed across more than one tissue, we observed a mean change in BMI of 0.015 kg/m^2^ per unit change in IOGC score (linear regression, *P* = 7×10^−7^), whereas increasing this threshold to >= 10 tissues results in a greater than 5-fold increase in mean change in BMI to 0.08 kg/m^2^ per unit change in IOGC score (linear regression, *P* = 0.04) (**Fig. 2G**). Using a permutation test of matched non-outlier variants (N permutations = 10,000), we again observed that outlier effects exceed that which would be expected using random subsets of rare variants (**Sup. Fig. 6B**).

We further investigated whether the severity of outlier gene expression integrated into the IOGC score affected change in BMI. We observed that at increasingly more-stringent Z-score thresholds, mean change in BMI also increased (**Fig. 2H**) (abs(Z) 2-3, linear regression *r* = 0.008 (*P* < 1×10^−6^); abs(Z) 3-4, linear regression *r* = 0.009 (*P* = 2×10^−6^); abs(Z) 4-5, linear regression *r* = 0.014 (*P* = 9×10^−4^). Comparing variants identified in outlier genes with Z-score between abs(Z) 2-3 with abs(Z) 4-5, we observe a 75% increase in mean change in BMI per unit change in IOGC score.

### Extreme IOGC scores lead to substantial deviation from PRS for BMI and obesity

From the analyses presented above, rare variants linked to outlier gene expression in GTEx had larger effects on BMI and rates of obesity, independent of PRS, and this effect is modulated by properties of outlier effects (i.e. multi-tissue outliers, outlier Z-score severity). We next sought to understand the magnitude of deviation from cohort-average BMI and obesity associated with outlier rare variant burden. For this analysis we used outlier-associated variants identified using the all-outlier method (**Methods**) - this increases the range of IOGC scores we can interrogate (range: top-outlier = −19:20; all-outliers = −67:69).

We calculated the mean rate of change in BMI across different percentiles of IOGC score using a linear regression model, adjusting for PRS, age, sex, first ten principal components of ancestry, and genotyping array. Each increment in IOGC score percentile bin is associated with a mean rate of change in BMI of 0.05 kg/m^2^ (linear regression, *P* < 1×10^−16^); comparing bottom and top 0.05% percentiles, this results in a difference in mean BMI of 0.74 kg/m^2^ (**Fig. 3A**). In the regression model, we tested for an interaction between PRS and IOGC score and observed no significant effect.

**Figure 3.**
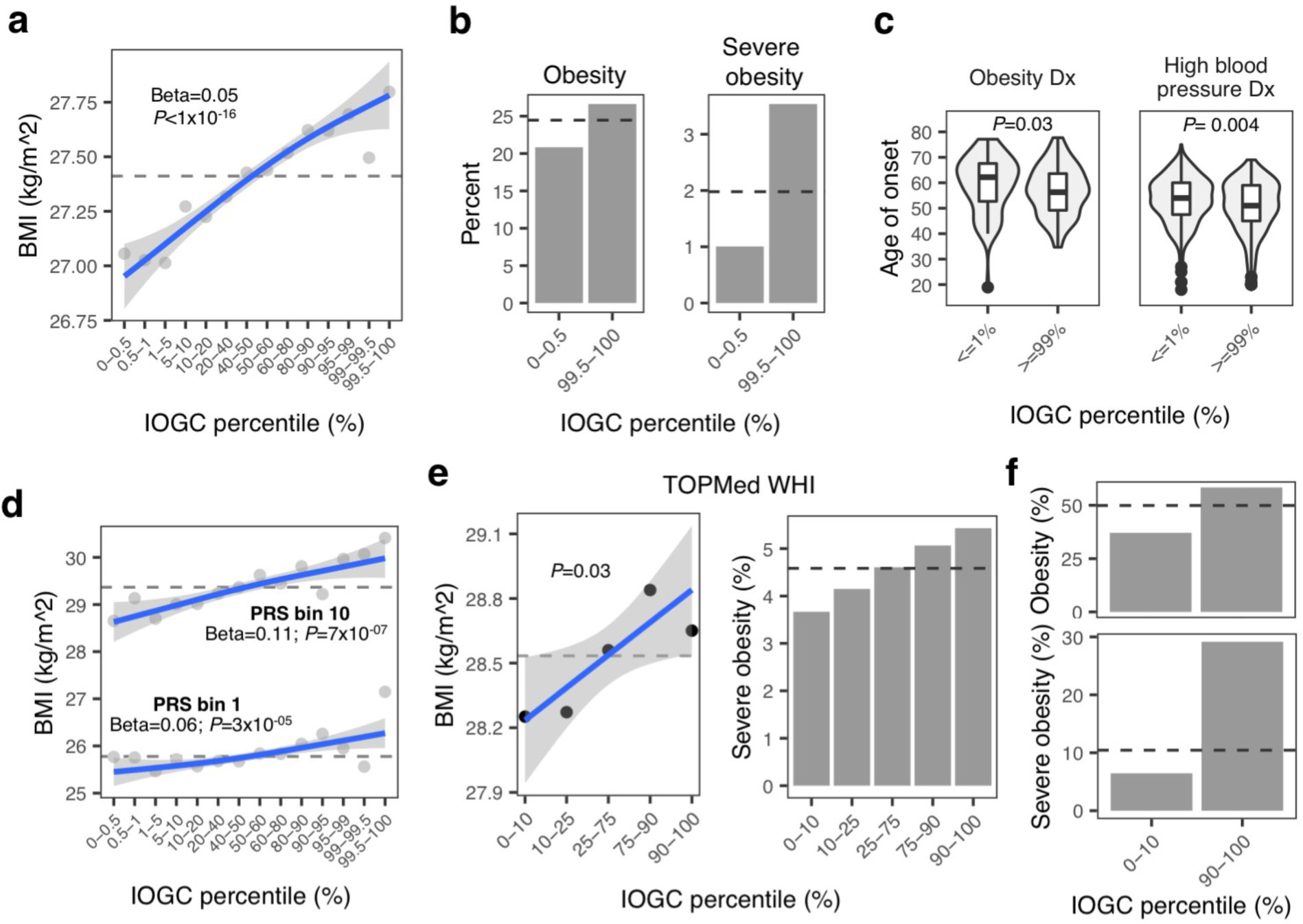
Increasing burden of outlier variants is associated with significant deviation in PRS-predicted body mass index and obesity. **A**. Mean BMI at different percentiles of IOGC score and linear regression fit. Dashed line indicates overall cohort mean. **B**. Rate of obesity and severe obesity for individuals with extreme IOGC scores (0.5% and 99.5% percentiles). Logistic regression results: obesity: 1.003 (*P* = 1.20×10^−14^); severe obesity: 1.004 (*P* = 0.0009). Dashed line indicates overall cohort mean. **C**. Age of onset of obesity and high blood pressure diagnosis for individuals with extreme IOGC scores. Obesity diagnosis: percentile <=1%: mean age of onset obesity = 59.41; percentile >=99%: mean age of onset 56.95 (*P* = 0.03), mean difference of 2.46 years; high blood pressure diagnosis: percentile <=1%: mean age of onset high blood pressure = 53.04; percentile >=99%: 50.44 (*P* = 0.004), mean difference of 2.6 years. **D**. Mean BMI at different percentiles of IOGC score, computed separately in PRS bins 1 and 10. Dashed line indicates the mean rate within each PRS group. **E**. Mean BMI (left) and incidence of severe obesity (right) at different percentiles of IOGC score, including linear regression fit, in TOPMed WHI. Dashed line indicates overall cohort mean. **F**. Mean incidence of obesity (top) and severe obesity (bottom) for TOPMed WHI PRS decile 10 cohort, using outlier variants with multi-tissue outlier count >=10. Dashed line indicates overall cohort mean for PRS decile.

Rates of obesity (BMI >= 30 kg/m^2^) and severe obesity (BMI >= 40 kg/m^2^) for individuals in the extreme 0.5% IOGC score percentiles deviate from the average rates of the full cohort overall: obesity: 0.5 percentile = 20.8%; 99.5 percentile = 26.6%; average overall: 24.5% (logistic regression, *P* = 5.3×10^−14^); severe obesity: 0.5 percentile = 1%; 99.5 percentile = 3.5%; average overall: 1.9% (logistic regression, *P* = 0.001) (**Fig. 3B**). We also tested for risk of being underweight (BMI<18.5 kg/m^2^) and found an inverse relationship (i.e. lower IOGC score percentile increases risk of being underweight) (logistic regression, *P* = 0.003).

Individuals in extreme IOGC score percentiles further differed in their age of onset of obesity and high blood pressure diagnosis (where high blood pressure is used as a proxy for hypertension). For diagnosis of obesity, individuals in IOGC score percentile <=1%, mean age of onset obesity = 59.41, whereas individuals in IOGC score percentile >=99%, mean age of onset = 56.95, a difference of in age of onset of 2.46 years (Wilcoxon test, *P* = 0.03). For high blood pressure diagnosis, individuals in IOGC score percentile <=1%, mean age of onset = 53.04, whereas individuals in IOGC score percentile >=99%, mean age of onset = 50.44, a difference of 2.6 years (Wilcoxon test, *P* = 0.004) (**Fig. 3C**).

We also observed that effects of outlier rare variants can manifest from childhood. We identified a subset of individuals in the UKB validation cohort (N=55,126) who provided self-reported information on being “plumper” or “thinner” than average at age 10 (UKB data field #1687). We tested the association of IOGC score with childhood body size using a logistic regression model (where the response was coded as 0=“thinner”, 1=“plumper”). The model was adjusted for PRS, age, sex, and first ten principal components of ancestry. For each unit change in IOGC score, we observed an increase in the odds of having a “plumper” comparative body size at 10 of 1.001 (logistic regression, *P* = 3×10^−09^).

We next sought to understand the potential magnitude of deviation in PRS-predicted rate of severe obesity (BMI >= 40 kg/m^2^) associated with extreme IOGC score. We first stratified individuals by PRS: “low-risk” (PRS decile 1); “high-risk” (PRS decile 10), and further subset by increasingly stringent percentiles of IOGC score (**Table 1**). We computed empirical P-values using a permutation test (N permutations = 10,000) to understand how likely the rates of severe obesity are observed across random subsets of individuals from within the same PRS groups. From this analysis we observed, for example, that “low-risk” individuals (PRS decile 1) in the 99^th^ percentile of IOGC score have a rate of severe obesity approaching the average rate for PRS decile 10 (4.55%, *P* = 0.0009), a greater than 6-fold increase in the PRS-predicted rate of severe obesity.

**Table 1.** Rates of severe obesity as a function of PRS and IOGC.

| IOGC percentile | $\leq 0.25\%$ | $\leq 0.5\%$ | $\leq 1\%$ | $\leq 10\%$ | PRS only | $\geq 90\%$ | $\geq 99\%$ | $\geq 99.5\%$ | $\geq 99.75\%$ |
| --- | --- | --- | --- | --- | --- | --- | --- | --- | --- |
| PRS bin 1 “low-risk” | 0%<br>(n: 45)<br>NS | 0%<br>(n: 84)<br>NS | 0%<br>(n: 171)<br>NS | 0.66%<br>(n: 1,351)<br>NS | 0.67%<br>(n: 12,094) | 1.01%<br>(n: 992)<br>NS | 4.55%<br>(n: 110)<br>$P=0.0009$ | 6.35%<br>(n: 63)<br>$P=0.0006$ | 6.06%<br>(n: 33)<br>$P=0.02$ |
| PRS bin 10 “high-risk” | 0%<br>(n: 27)<br>NS | 1.72%<br>(n: 58)<br>NS | 2.65%<br>(n: 113)<br>NS | 4.08%<br>(n: 1152)<br>NS | 4.99%<br>(n: 12,043) | 5.28%<br>(n: 1137)<br>NS | 7.63%<br>(n: 118)<br>NS | 10.35%<br>(n: 58)<br>NS | 16.67%<br>(n: 30)<br>$P=0.02$ |

\* P-values are calculated empirically across 10,000 permutations

We performed regression modelling within the “low-risk” and “high-risk” PRS groups described above, and observed similar linear regression coefficients per change in IOGC score percentile bin (linear regression; PRS bin 1: *r* = 0.06 (*P* = 3×10^−5^); PRS bin 10: *r* = 0.11 (*P* = 7×10^−7^) (**Fig. 3D**). We next investigated the composition of outlier variants among individuals with outlier IOGC scores (specifically, bottom and top 10% of IOGC score distribution). For each individual in this set, we calculated the IOGC-aware deviation (i.e. below mean for low IOGC individuals, and above mean for high IOGC individuals) from within-group PRS mean BMI (as Z-score) and looked for differences in the relative composition of outlier variants in individuals near the mean (abs(Z-score)>0 and <0.5) and far from mean (abs(Z-score)>=3). We observed that individuals far from their predicted PRS mean were enriched for missense and splicing outlier variants (Fisher’s Exact Test; missense: odds ratio = 1.27 (CI 1.14 – 1.40), *P* = 8×10^−6^; splicing: odds ratio = 1.25 (CI 0.98 – 1.58), *P* = 0.05) (**Sup. Fig. 7**). This finding suggests that variant annotations could be integrated in to future iterations of IOGC to further increase the predictive power of the score. Furthermore, we computed IOGC scores across genes summarized by their effect in the PRS (using the maximum effect weight per PRS variant mapped to within each gene), observing that IOGC score increases as a function of gene PRS effect size (**Sup. Fig. 8**). This results demonstrates that IOGC score is increased in genes with known larger effects on BMI.

### Replication in TOPMed WHI

We replicated our findings using TOPMed WHI data, subset to individuals with European ancestry and with genetic and phenotypic data available (N = 6,501). We constructed a linear regression model including PRS, first ten principal components of ancestry, age, and IOGC score (sex is not included since TOPMed WHI is an all-female cohort). Variant effect directions were obtained from UKB GWAS, as above. IOGC score is again a significant predictor of BMI (mean change in BMI per quantile of IOGC score: linear regression *r* = 0.13 kg/m^2^, *P* = 0.03) (**Fig. 3E**). Although explicitly tested in the regression model, we visually compared the PRS of individuals in the 10th and 90th percentile of IOGC score and observed no significant differences in PRS for these two groups (**Sup. Fig. 9**). In the regression model, we again tested for an interaction between PRS and IOGC score and observed no significant effect.

Similar to observations in UKB, rates of obesity (BMI >= 30 kg/m^2^) and severe obesity (BMI >= 40 kg/m^2^) for individuals in the 10^th^ and 90^th^ percentiles for IOGC score deviated from the average rates of the full cohort overall: obesity: 10^th^ percentile = 31.03%; 90^th^ percentile = 35.30%; average overall: 33.49% (**Sup. Fig. 10**); severe obesity: 10^th^ percentile = 3.67%; 90^th^ percentile = 5.43%; average overall: 4.59% (**Fig. 3E**).

Subsetting by multi-tissue outlier-associated variants (N tissues >= 10), we again observed a significant effect of IOGC score independent from PRS, age and genetic ancestry (mean change in BMI per quantile of IOGC score: linear regression *r* = 0.20 kg/m^2^, *P* = 0.01). Further highlighting the independence of IOGC score from PRS, risk of obesity and severe obesity among individuals within PRS decile 10 (“high-risk”) can vary substantially from average for individuals in the 10^th^ and 90^th^ percentile of IOGC score (**Fig. 3F**).

### Rare variants impact PRS prediction across multiple traits and diseases

The main focus of our study was on BMI and associated rates of obesity, but the same approach can be applied to many other traits and diseases. For example, using a publicly-available PRS for type-2 diabetes (T2D, **Methods**), we observed a deviation from PRS-predicted mean incidence of diabetes associated with an increasing burden of outlier variants (IOGC *r* = 1.01, *P* = 0.03, logistic regression) (**Fig. 4A**). Looking at age of T2D onset in a cohort defined as “high-risk” by PRS (PRS Z-score > 1), we observe a difference in mean age of onset of 4.04 years (Wilcoxon test, *P* = 0.02), comparing individuals in the 10^th^ and 90^th^ percentiles of IOGC score among this PRS high-risk group (**Fig. 4B**).

**Figure 4.**
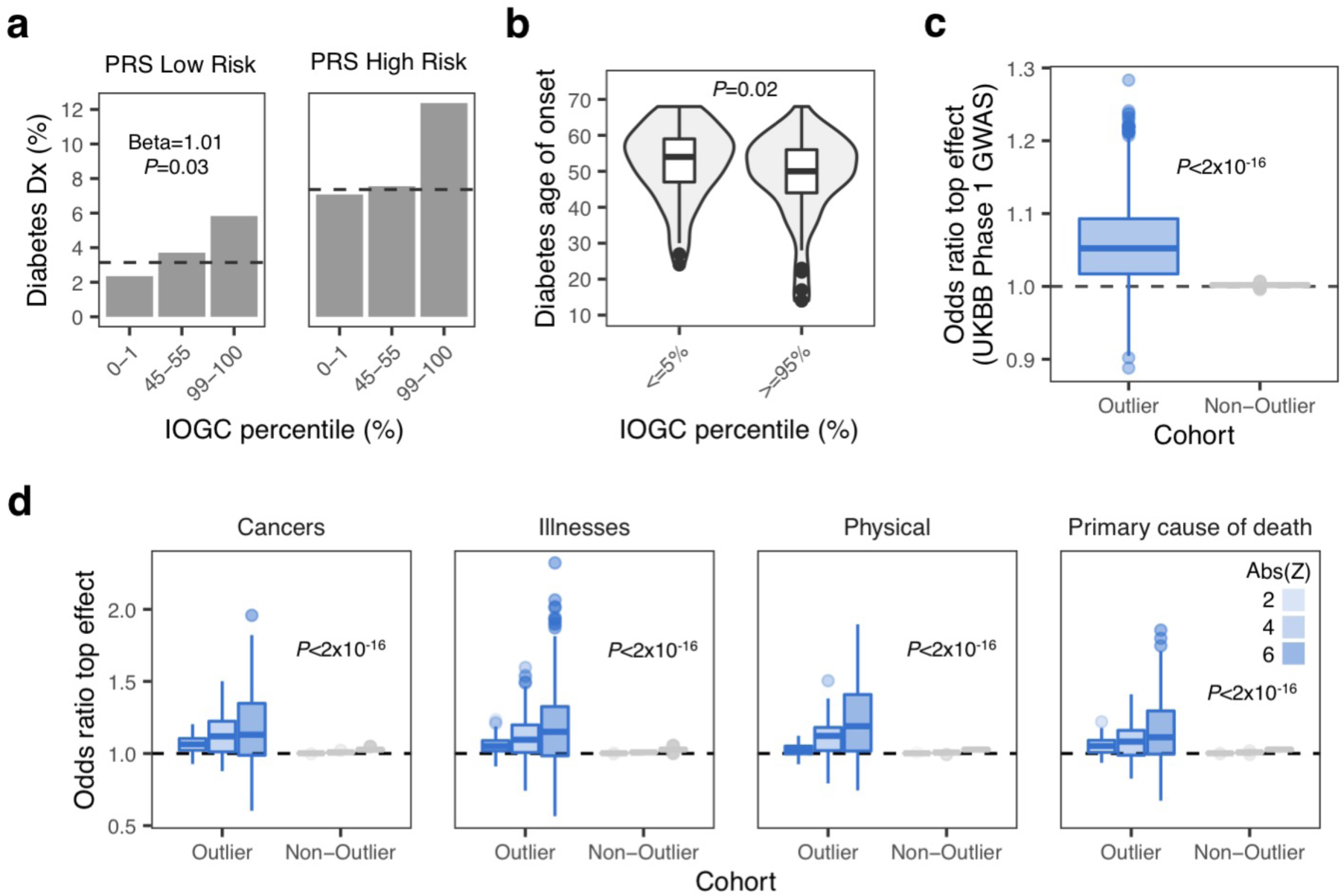
Extending the method to diverse traits and diseases. **A**. Deviation from PRS-predicted mean incidence of diabetes (%) amongst individuals with extreme IOGC scores, and average score. Logistic regression: beta = 1.01 (*P* = 0.03). Dashed line shows the average incidence of diabetes for each PRS bin. Low-risk = PRS bin 1 of 5 (PRS Z-score < −0.84); High-risk = PRS bin 5 of 5 (PRS Z-score > 0.84). **B**. Age on onset of diabetes in PRS z-score > 1 cohort: difference in mean age of onset = 4.04 years (Wilcoxon test, *P* = 0.02). **C**. Distribution of mean odds ratio per UKB GWAS phenotype across 1,000 permutations for outlier-vs. non-outlier associated variants (blue) and non-outlier vs. non-outlier variants (gray) (Wilcoxon test, P<2×10^−16^). **D**. Distribution of mean odds ratio per UKB GWAS phenotype (meta-groups: cancer; illnesses; physical; primary cause of death) across 1,000 permutations for outlier-vs. non-outlier associated variants (blue) and non-outlier vs. non-outlier associated variants. Analysis was repeated at increasing thresholds of outlier gene expression absolute Z-score (from abs(Z-score)>=2 to abs(Z-score)>=6).

To quantify differences in effect sizes of outlier-associated variants across diverse traits and disease, we repeated the same permutation test described earlier (**Methods**), utilizing all outlier- and non-outlier variants identified using the “top-outlier” method across 2,419 traits and diseases released in UKB Phase 1 GWAS (N permutations = 1,000). We observed a mean odds ratio of 1.05 (SD = 0.06) across all disease and traits when comparing outlier vs. non-outlier variants, and a mean odds ratio of 1 (SD = 0.002) for non-outlier variants only (Wilcoxon test, *P* < 2×10^−16^) (**Fig. 4C**). Increasing the outlier Z-score threshold, we observed an increasing trend for observing outlier variants with top effect sizes (mean odds (SD): abs(Z-score)>4 = 1.10 (0.14); abs(Z-score)>6 = 1.17 (0.25) (Wilcoxon test, *P* < 1×10^−16^ both comparisons). No difference was observed in odds ratios when comparing non-outlier variants only.

Across different GWAS meta-categories (cancer, illnesses, physical traits, cause of death), we observed the same overall trend for observing outlier variants with top GWAS effect sizes, increasing with Z-score threshold (*P* < 2×10^−16^ for all outlier vs. non-outlier comparisons) (**Fig. 4D**). For example, for breast cancer (ICD-10: C50, Malignant neoplasms of breast), we observed odds ratios 1.02, 1.11, 1.25 for abs(Z-score)>2, 4 and 6, respectively. We also expected some GWAS traits to not be sensitive to SNPs linked to gene outlier effects. We explored this hypothesis by manually selecting several traits where genetic relationships are more speculative (i.e. loud music exposure, transport for commuting to job) and observed no difference in the distribution of odds ratios comparing outlier vs. non-outlier variants and non-outlier variants only (**Sup. Fig. 11**).

## Discussion

Integration of rare variants within polygenic risk predictions is a major challenge. We have demonstrated that a high burden of rare variants can lead to substantial deviations in PRS-predicted phenotype. Furthermore, by integrating rare variants into genetic risk prediction using the IOGC score, we demonstrate improvements in predicting risk for multiple traits and diseases. Specifically, for BMI we demonstrate that PRS “low-risk” individuals who are in the top 1% of IOGC have a rate of severe obesity (BMI>=40 kg/m^2^) approaching the average rate for PRS “high-risk” individuals. By leveraging diverse traits and diseases recorded in the UKB we further demonstrate applications for diverse polygenic phenotypes.

Notably, the power of this approach is enabled by first isolating rare, expression outlier-linked variants in GTEx. Given that this cohort is limited to 546 individuals, it is certain that many large-effect variants impacting expression remain to be identified. Future large-scale RNA-sequencing studies and catalogues of outlier-associated rare variants will only increase the efficacy of this approach. Furthermore, we could recover only a subset of outlier-associated rare variants in UKB, due to limitations in imputation; future WGS in population biobanks will recover more rare, outlier-associated variants. Future WGS will also expand the frequency spectra that can be interrogated - carriers of ultra-rare, outlier variants are likely to have even larger effects and impacts on the IOGC score^19–21^. Additionally, future work could integrate other data modalities (e.g. single cell, proteomics outliers).

Our study offers a baseline of phenotypic effects of rare, large-effect variants and shows considerable impact in aiding the prediction of individual phenotypes. As with current genetic risk prediction, we expect that the IOGC score can immediately help to better identify and stratify high-risk individuals into specific early treatments. Overall, this work has important immediate implications for the implementation of genetic risk prediction in standard clinical care.

## Methods

### GTEx v7 data

Processed WGS variant data was obtained from GTEx v7 (see **Resource Availability**). Using the software bedtools^22^ (--window flag), variants were linked to genes if falling within the gene body, 10 Kb upstream of transcription start start, or 10 Kb downstream of the transcription end site. Using the software Vcfanno^23^, SNP variants were intersected with gnomAD (version r2.0.2)^17^ and CADD^18^ databases to obtain the minor allele frequency and CADD score, respectively, for each variant. Variant annotations were obtained using Variant Effect Predictor (VEP) (version 88)^24^. Minor allele frequencies were calculated across all individuals in gnomAD. Non-SNP (i.e. indel, SNV) variants were discarded. SNPs were retained if the gnomAD MAF fell in the range 0<MAF<1%. Multi-allelic SNPs were removed (multi-allelic in gnomAD). Finally, SNPs were required to have been directly measured or imputed in UKB Phase 1 GWAS (imputation quality as described in [^25^], namely: UKB MAF > 0.1%; Hardy-Weinberg Equilibrium P > 1×10^−10^; INFO score > 0.8).

Processed RNA-sequencing data was obtained from GTEx v7 (see **Resource Availability**). To identify GTEx outlier gene expression samples, normalized gene expression values (FPKM) were processed across all GTEx v7 tissues, limited to autosomal genes annotated as protein coding or long non-coding RNA genes in GENCODE v19. A minimum expression filter was applied per gene (>=10 individuals with FPKM > 0.1 and read count > 6); genes not passing this filter were removed. Expression values were PEER^26^ factor corrected (using 15 factors for tissues with <= 150 samples, 30 for tissues with <= 250 samples, and 35 for tissues with > 250 samples), then scaled and centered to generate expression Z-scores. Individuals exhibiting global patterns of outlier gene expression for a given tissue were removed from the final corrected expression matrix for that tissue. Global outlier is defined as any individual who has the most-extreme absolute Z-score of corrected gene expression in 100 or more genes in a given tissue at an outlier cutoff of abs(Z-score)>2.

### UK Biobank data

UKB Phase 1 GWAS summary statistics were downloaded from the Neale Lab server (available at http://www.nealelab.is/uk-biobank). Summary statistics for each GTEx outlier and non-outlier variant were joined on chromosome, position, ref, and alt columns, using hg19 coordinates. All other phenotypic and genotypic data were sourced from the data instance approved under UKB application #24983 (see **Resource Availability**). Individual-level phenotypes for weight (UKB data field #21002), body mass index (UKB data field #21001) and diabetes (UKB data field #2443) were downloaded from the relevant phenotype file. For weight and BMI, we averaged (using the median) over all observations per-individual for those individuals with multiple observations for the same phenotype. Imputed and directly measured genotypes for all variants used in this study were extracted from the genotyping callset (version 3). Additional phenotypic and demographic data used included: age, sex, principal components, genotyping array (all included in the UKB sample QC file); age of onset of diagnosis (obesity (UKB data field #130792), high blood pressure (UKB data field #2966), diabetes (UKB data field #2976)); and comparative body size at age 10 (UKB data field #1687).

### TOPMed Women’s Health Initiative (WHI) data

The full TOPMed WHI cohort was first subset to self-reported European ancestry only (race code ‘5’ in file WHI.phv00078450.v6.p3.c1.txt). Individual-level weight and BMI measurements were obtained from the file phs000200.v11.pht001019.v6.p3.c1.f80_rel1.HMB-IRB.txt.gz. The average (median) was found for individuals with multiple observations of the same phenotype. Genotypes were obtained from whole genome sequencing data available in the archive phg001146.v1.TOPMed_WGS_WHI.genotype-calls-vcf.c1.HMB-IRB.tar. BED files were created using the software plink (version 2.0)^27^. TOPMed WHI bed files are in hg38 assembly; we used the software CrossMap^28^ to convert genome coordinates from hg19 to hg38 assemblies for the purposes of measuring GTEx outlier and non-outlier variants among TOPMed WHI individuals and computing polygenic risk scores.

Genotype principal components we computed using a random selection of common variants (N=50,000) available in UKB; we chose to leverage UK Biobank allele count information to define a set of high-confidence common variants (UKB minor allele count > 50,000), given the increased sample size of UKB compared with TOPMed WHI. Genotypes were extracted using plink (version 2.0). To create the input matrix for computed principal components, the genotypes of each extracted variant was imported and checked for minor allele variants and the percentage of missing genotypes; alles with zero minor allele variants and/or >1% missingness were removed. For variants with >0 and <=1% genotype missingness, missing genotypes were replaced by the mode for that particular variant. Principal components were computed using the software flashpca^29^.

### Million Veteran Program (MVP) GWAS for body mass index

DNA extracted from participants’ blood was genotyped using a customized Affymetrix Axiom® biobank array, the MVP 1.0 Genotyping Array. The array was enriched for both common and rare genetic variants of clinical significance in different ethnic backgrounds. Quality-control procedures used to assign ancestry, remove low-quality samples and variants, and perform genotype imputation to the 1000 Genomes reference panel were previously described^30^. Individuals related more than second degree cousins were excluded.

We recently conducted HARE (Harmonized ancestry and Race/Ethnicity) analysis using race/ethnicity information from MVP participants^31^. Genotyped MVP participants are assigned into one of the four HARE groups (Hispanics, non-Hispanics White, non-Hispanics Black, and non-Hispanics Asian) and “Other”. The analysis is based on a machine learning algorithm, which integrates race/ethnicity information from MVP baseline survey and high-density genetic variation data. Trans-ethnic, and ethnicity-specific principal component analyses were performed using flashPCA^29^. BMI was calculated as average BMI using all measurements within a three-year window around the date of MVP enrollment (i.e., 1.5 years before/after the date of enrollment), excluding height measurements that were >3 inches or weight measurements >60 pounds from the average of each participant.

Genetic association with BMI in the MVP cohort was examined among 217,980 non-Hispanic White participants. BMI was stratified by sex and adjusted for age, age-squared, and the top ten genotype-derived principal components in a linear regression model. The resulting residuals were transformed to approximate normality using inverse normal scores. Imputed and directly measured genetic variants were tested for association with the inverse normal transformed residuals of BMI through linear regression assuming an additive genetic model.

### Isolating rare variants observed in GTEx gene expression outliers and non-outliers

Rare variants occurring in gene expression outlier individuals are identified using two methods; namely, top-outlier and all-outliers. Both approaches start with genetic and transcriptomic data processed as detailed above (“GTEx v7 genetic and transcriptomic data”). Using the top-outlier approach, variants were aggregated across all individuals and subset to gnomAD MAF > 0 and < 1%. This list was then tabulated to obtain a count of unique individuals with each variant; any variant observed in > 1 individual was removed. As a further filtering step, variants were retained only if they were included in UKB Phase 1 GWAS. To link variants to expression outliers, we identified for each tissue the individuals with the least or most expression per gene (i.e. under-expression outlier and over-expression outlier), removing any results falling below a predefined Z-score threshold of abs(Z-score) < 2. We also defined, for each tissue and gene, a set of individuals with non-outlier gene expression (defined as abs(Z-score)<1). Non-outlier variants were filtered to match the CADD score (within a window +/- 5) of any outlier variants for each tissue/gene/outlier direction triple; this is important for genes with both an under-expression and over-expression outlier, as subsequent permutation testing uses outlier and non-outlier variants matched on a CADD score window. Variants identified in outliers in >=1 tissue were ignored when identifying matching non-outlier variants; in this way, a putatively causal large-effect expression variant (in any number of tissues) would not be counted in both outlier and non-outlier variant sets. For the all-outliers method, we removed any outlier variant also identified in >= 1 non-outlier individual; this differs from the top-outlier method, in which outlier variants identified in any other individual (regardless of outlier status) are removed. We did not define a matching set of non-outlier variants in the all-outliers method, due to run-time constraints on the computational pipeline developed for this study. For both methods, we recorded the number of tissues in which each outlier variant was identified (for variants identified in >1 tissue, we refer to these as multi-tissue outlier variants).

### GWAS effect size permutation test

We performed a permutation test to study differences in GWAS effect sizes for GTEx outlier and non-outlier variants. This test was repeated for two independent GWAS cohorts: UK Biobank and Million Veteran Program. For each GWAS, the input data is a file containing outlier and non-outlier variants with associated GWAS effect size (i.e. beta estimate), linked outlier gene, GTEx sample ID, outlier direction (under-expression/over-expression), and outlier tissue. Additionally, for GWAS of traits and disease where we also run a separate test after integrating PRS information, subsetting genes to those linked to any outlier variant falling within 10 Kb of a PRS variant. To define a set of outlier and non-outlier variants, we first subset outlier variants using a defined absolute Z-score of outlier gene expression, then find the intersection (using tissue, gene, and outlier direction) between the outlier variants that pass the Z-score thresholds and the matched non-outlier controls. This step ensures we have sufficient data to randomly select exactly one outlier and non-outlier variant per tissue/gene/outlier direction triple (we refer to this as an outlier triple), per permutation. The permutation test is based on the results of the top-outlier methods (see previous section); therefore, there is exactly one outlier individual per outlier triple. However, for non-outlier variants, there can be matched variants identified in > 1 unique individual. We subset randomly to one non-outlier individual per outlier triple, then randomly select exactly one outlier and non-outlier variant per outlier triple. For each outlier triple, we then count which variant is associated with the greater GWAS absolute effect size (outlier/non-outlier); this information can then be summarized in a contingency table, which is then used as the input to compute an odds ratio. We repeated this analysis for a file containing non-outlier variants only, which follows the same method described above, comparing two randomly chosen non-outlier variants per outlier triple, for outlier triples with non-outlier variants from >= 2 unique non-outlier individuals.

### Calculating polygenic risk scores

We computed polygenic risk scores (PRS) for the UKB and TOPMed WHI cohorts in this study. Two high-quality, publicly-available PRS were used (see **Resource Availability**): body mass index (Khera.et.al_GPS_BMI_Cell_2019.txt.zip); and type-2 diabetes (Type2Diabetes_PRS_LDpred_rho0.01_v3.txt). Scores were calculated using the software plink (version 2.0) (--score flag). PRS variant coordinates were first converted to hg38 assembly using CrossMap^28^ for calculating PRS scores in the TOPMed WHI cohort. Scores were calculated separately for each chromosome, then summed per individual and scaled to generate Z-scores.

### GTEx eQTL to assess concordance in GWAS effect direction between eQTL and outlier variants

GTEx eQTL summary statistics were downloaded and filtered on P-value (using column “pval_nominal”) with threshold P<1×10^−18^. Remaining variants were linked to their GWAS effect size (i.e. protective or risk). For genes with >1 eQTL passing the P-value threshold, the variant with the smallest UKB GWAS P-value was retained. This step was computed separately for each GTEx tissue. We used a majority-rule approach to assign a single, high-quality consensus GWAS effect direction per gene, based on the median slope estimate and GWAS effect direction across all top eQTL variants per tissue that passed the two P-value filtering steps. For example, if a given gene consisted of 10 eQTL risk variants and 5 protective risk variants, the gene would be assigned a “risk” label. We removed any genes where a majority GWAS effect direction could not be computed (i.e. an equal number of protective and risk effects), genes where eQTL risk variants shared a median slope that matched the median slope of eQTL protective variants (e.g. positive slope in both cases), and genes where the slope of any eQTL risk variant matches the slope of any eQTL protective variant. Outlier variants are then compared on the slope and GWAS effect direction of the consensus eQTL results (e.g. for a given gene with positive median slope and GWAS risk effect, we assessed if the outlier variant was an over-expression outlier variant and its comparable relationship to GWAS risk).

### Inferring UKB non-British white validation cohort

Using the self-identified non-British white labels that were reported in the UKB metadata, a larger cohort of predicted non-British white individuals was inferred. For all self-reported non-British white individuals, the mean and standard deviation of the first and second genotypic principal components were calculated. All individuals without a self-reported ethnic identity that were within +/- 3 SD of the calculated mean PC1 and PC2 values were inferred to be non-British white. All self-reported non-British white individuals that fell out of this range were also excluded. This final cohort consisted of 23,790 self-reported non-British individuals, and 97,154 inferred non-British white individuals. We found that the PRS distribution of this non-British white cohort did not differ significantly from a normal distribution (Shapiro-Wilk normality test; *P* = 0.2774), suggesting that the PRS as calculated on the British white cohort generalizes well to this cohort.

### Quantifying effect on phenotypes associated with IOGC score

Using the list of GTEx outlier variants linked to genes, we retained the genes in which >= 1 outlier variant overlapped any PRS variants (within a +/- 10 Kb window). In this way, we focus only on genes previously linked to the phenotype (and therefore included in the PRS). The resulting set of outlier- and non-outlier variants in retained genes were written to a lookup file which was then input to the software plink^27^ (--extract flag) to identify UKB individuals in the validation cohort who are heterozygous or homozygous for each variant (i.e. alternate allele genotype 1 and 2, respectively). We then used previously-released UKB GWAS effect estimates to assign effect directions to each outlier variant (i.e. risk/protective). Given that the previously-released GWAS were calculated on UKB individuals with white British genetic ancestry (see^25^), the non-British white cohort validation cohort we constructed for this study, as well as the TOPMed WHI cohort, was non-overlapping.

We quantified the effect of outlier variant burden on phenotype by computing a score that summarizes, per individual, putative outlier gene burden. We refer to this quantity as the independent outlier gene count (IOGC). To compute this score, for each individual we link variants to effect size direction in UKB, then collapse to gene-level to prevent double-counting. Per individual, we convert the beta effect estimate per variant to integers using a sign function:

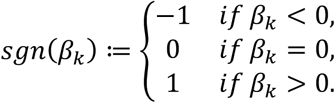

where *β* is the UKB GWAS beta coefficient for variant *k*. In practice, effect sizes of zero are not generally observed, so we expect to see only values of −1 or 1. Following this step, we take the distinct values per gene (i.e. remove duplicates); since our goal is to use outlier variants to tag outlier/dysregulated gene expression, this step prevents counting of putative outlier gene expression more than once. Therefore, if we denote the vector of *sgn* (*β*_*k*_) for variants linked to a given gene as *s*, then:

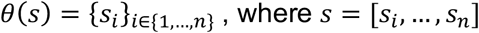

This is repeated across all genes *g* linked to >=1 outlier variant, and summed to yield the IOGC score for each individual *j*:

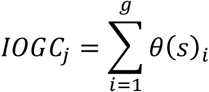

Linear regression was used for quantitative phenotypes, and logistic regression for binary phenotypes. In the regression models, we adjusted for PRS, age, sex (UKB only since TOPMed WHI is a female-only cohort), first ten principal components of genetic ancestry, and genotyping array (UKB only).

All statistical analyses were performed using R (version 3.6.0). Plots were generated using ggplot2 (version 3.3.0)^32^.

## Data Availability

GTEx (v7) RNA-seq and WGS data is available from dbGaP (dbGaP Accession phs000424.v7.p2)
GTEx (v7) eQTL summary statistics were downloaded from the GTEx Portal available at https://gtexportal.org/home/datasets
Data from the TOPMed Women's Health Initiative is available from dbGaP (dbGaP Accession phs001237)
UK Biobank (UKB) data was obtained under application number 24983 (PI: Dr. Manuel Rivas)
UKB Phase 1 GWAS summary statistics were downloaded from the Neale Lab server available at http://www.nealelab.is/uk-biobank
Polygenic risk scores (PRS) for body mass index and type-2 diabetes were downloaded from the Cardiovascular Disease Knowledge Portal available at http://kp4cd.org/dataset_downloads/mi
Gene annotation data was obtained from GENCODE (version 19) available at https://www.gencodegenes.org/human/release_19.html
Allele frequency data was obtained from gnomAD (version r2.0.2) available at https://console.cloud.google.com/storage/browser/gnomad-public/release/2.0.2/
hg19 coordinates were converted to hg38 using the chain file available at http://hgdownload.soe.ucsc.edu/goldenPath/hg19/liftOver/ Custom scripts to conduct all analyses not performed using existing software can be found at https://github.com/csmail/outlier_prs

## Resource availability

GTEx (v7) RNA-seq and WGS data is available from dbGaP (dbGaP Accession phs000424.v7.p2)

GTEx (v7) eQTL summary statistics were downloaded from the GTEx Portal available at https://gtexportal.org/home/datasets

Data from the TOPMed Women’s Health Initiative is available from dbGaP (dbGaP Accession phs001237)

UK Biobank (UKB) data was obtained under application number 24983 (PI: Dr. Manuel Rivas)

UKB Phase 1 GWAS summary statistics were downloaded from the Neale Lab server available at http://www.nealelab.is/uk-biobank

Polygenic risk scores (PRS) for body mass index and type-2 diabetes were downloaded from the Cardiovascular Disease Knowledge Portal available at http://kp4cd.org/dataset_downloads/mi

Gene annotation data was obtained from GENCODE (version 19) available at https://www.gencodegenes.org/human/release_19.html

Allele frequency data was obtained from gnomAD (version r2.0.2) available at https://console.cloud.google.com/storage/browser/gnomad-public/release/2.0.2/

hg19 coordinates were converted to hg38 using the chain file available at http://hgdownload.soe.ucsc.edu/goldenPath/hg19/liftOver/

Custom scripts to conduct all analyses not performed using existing software can be found at https://github.com/csmail/outlier_prs

## Acknowledgements

CS is supported by NIH grant T32LM012409. NMF is supported by a National Science Foundation Graduate Research Fellowship (grant number DGE 1656518) and a graduate fellowship from the Stanford Center for Computational, Evolutionary and Human Genomics. MGD is supported by a National Science Foundation Graduate Research Fellowship. MA is supported by the National Library of Medicine under training grant T15LM007033. XL is supported by the National Natural Science Foundation of China (grant number 31970554), National Key R&D Program of China (grant number 2019YFC1315804) and Shanghai Municipal Science and Technology Major Project (grant number 2017SHZDZX01). MJG is supported by a Stanford Graduate Fellowship. MAR is partially supported by Stanford University and a National Institute of Health center for Multi- and Trans-ethnic Mapping of Mendelian and Complex Diseases grant (5U01 HG009080) and partially supported by the National Human Genome Research Institute (NHGRI) of the National Institutes of Health (NIH) under award R01HG010140. SBM is supported by NIH grants U01HG009431, R01HL142015, R01HG008150, R01AG066490 and U01HG009080. This research has been conducted using the UK Biobank Resource under Application Number 24983, “Generating effective therapeutic hypotheses from genomic and hospital linkage data” (http://www.ukbiobank.ac.uk/wp-content/uploads/2017/06/24983-Dr-Manuel-Rivas.pdf). Based on the information provided in Protocol 44532 the Stanford IRB has determined that the research does not involve human subjects as defined in 45 CFR 46.102(f) or 21 CFR 50.3(g). All participants of UK Biobank provided written informed consent. This work in-part used supercomputing resources provided by the Stanford Genetics Bioinformatics Service Center, supported by National Institutes of Health S10 Instrumentation Grant S10OD023452. The content is solely the responsibility of the authors and does not necessarily represent the official views of the National Institutes of Health. The WHI program is funded by the National Heart, Lung, and Blood Institute, National Institutes of Health, U.S. Department of Health and Human Services through contracts HHSN268201600018C, HHSN268201600001C, HHSN268201600002C, HHSN268201600003C, and HHSN268201600004C. Molecular data for the Trans-Omics in Precision Medicine (TOPMed) program was supported by the National Heart, Lung and Blood Institute (NHLBI). See the TOPMed Omics Support Table below for study specific omics support information. Core support including centralized genomic read mapping and genotype calling, along with variant quality metrics and filtering were provided by the TOPMed Informatics Research Center (3R01HL-117626-02S1; contract HHSN268201800002I). Core support including phenotype harmonization, data management, sample-identity QC, and general program coordination were provided by the TOPMed Data Coordinating Center (R01HL-120393; U01HL-120393; contract HHSN268201800001I). We gratefully acknowledge the studies and participants who provided biological samples and data for TOPMed. This research is also supported by funding from the Department of Veterans Affairs Office of Research and Development, Million Veteran Program (MVP) Grant I01-BX003340 and I01-BX003362. This publication does not represent the views of the Department of Veterans Affairs or the United States Government. A list of MVP investigators can be found in supplementary materials. The funders had no role in study design, data collection and analysis, decision to publish, or preparation of the manuscript.

The authors would like to thank Jonathan Pritchard, Hakhamanesh Mostafavi, and members of the Pritchard Lab at Stanford University for helpful comments on this manuscript.

## TOPMed Omics Support Table

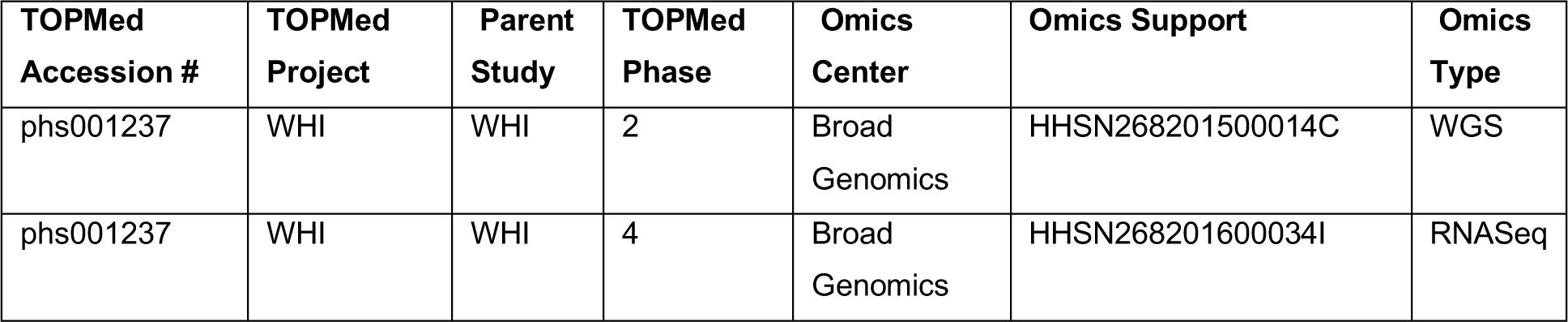

| TOPMed Accession # | TOPMed Project | Parent Study | TOPMed Phase | Omics Center | Omics Support | Omics Type |
| --- | --- | --- | --- | --- | --- | --- |
| phs001237 | WHI | WHI | 2 | Broad Genomics | HHSN268201500014C | WGS |
| phs001237 | WHI | WHI | 4 | Broad Genomics | HHSN268201600034I | RNASeq |

## Author contributions

Conceptualization, C.S. and S.B.M.; Methodology, C.S. and S.B.M.; Software, C.S. and N.M.F.; Formal Analysis, C.S., N.M.F., M.G.D., A.S.R., X.L., and M.J.G.; Investigation, C.S., N.M.F., M.G.D., A.S.R., and S.B.M.; Data Curation, T.L.A., M.A., and M.A.R.; Resources, Q.H., J.H., T.L.A., C.J.O., Y.V.S., C.K., A.R., and M.A.R.; Writing - Original Draft, C.S. and S.B.M.; Writing - Review & Editing, C.S., N.M.F., M.G.D., A.S.R., M.A., Q.H., J.H., T.L.A., C.J.O., Y.V.S., M.A.R., C.K., A.R., and S.B.M.; Funding Acquisition, M.A.R. and S.B.M.

