## Supplemental figures for "Integration of rare large-effect expression variants improves polygenic risk prediction"

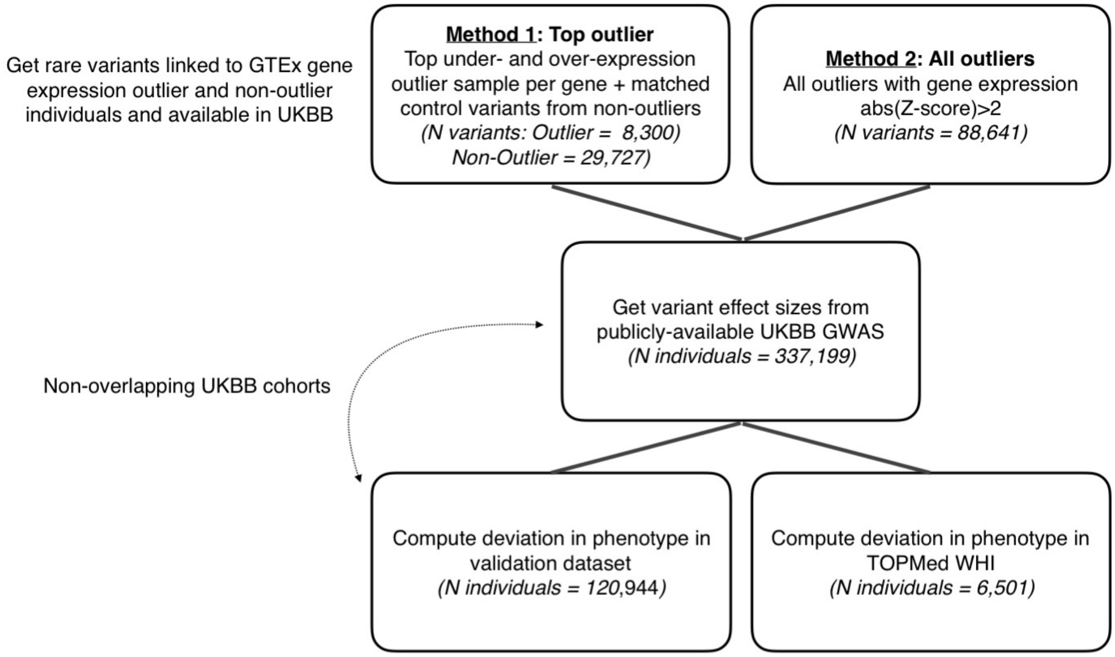


**Supplementary Figure 1.** Variant count for “top-outlier” and “all-outliers” methods, and sample sizes of UK Biobank and TOPMed WHI cohorts used in the study.

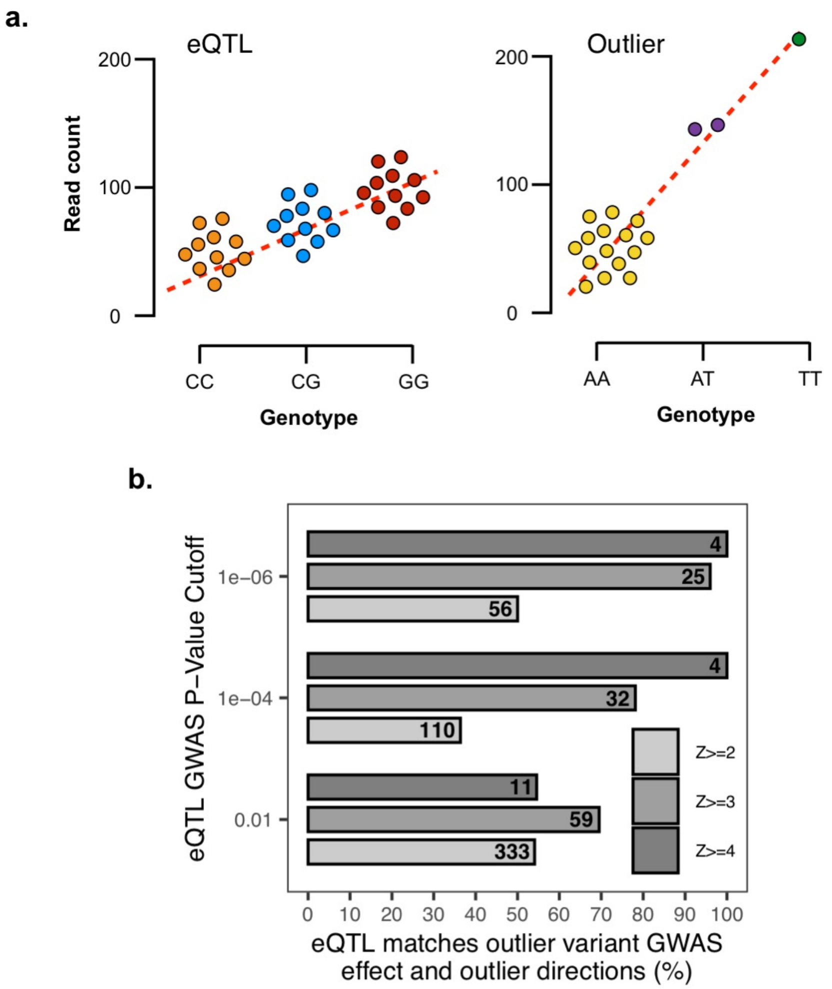


**Supplementary Figure 2. A.** Leveraging GTEx (v7) eQTL summary statistics to assess concordance in eQTL slope and outlier variant direction (i.e. under-expression or over-expression outlier) and GWAS effect direction. **B.** Rate of concordance increases with eQTL GWAS P-value cutoff and outlier variant Z-score threshold. Z-score bins collapsed to abs(Z-score) for purposes of visualization. Numbers indicate the total number of variants at the indicated thresholds.

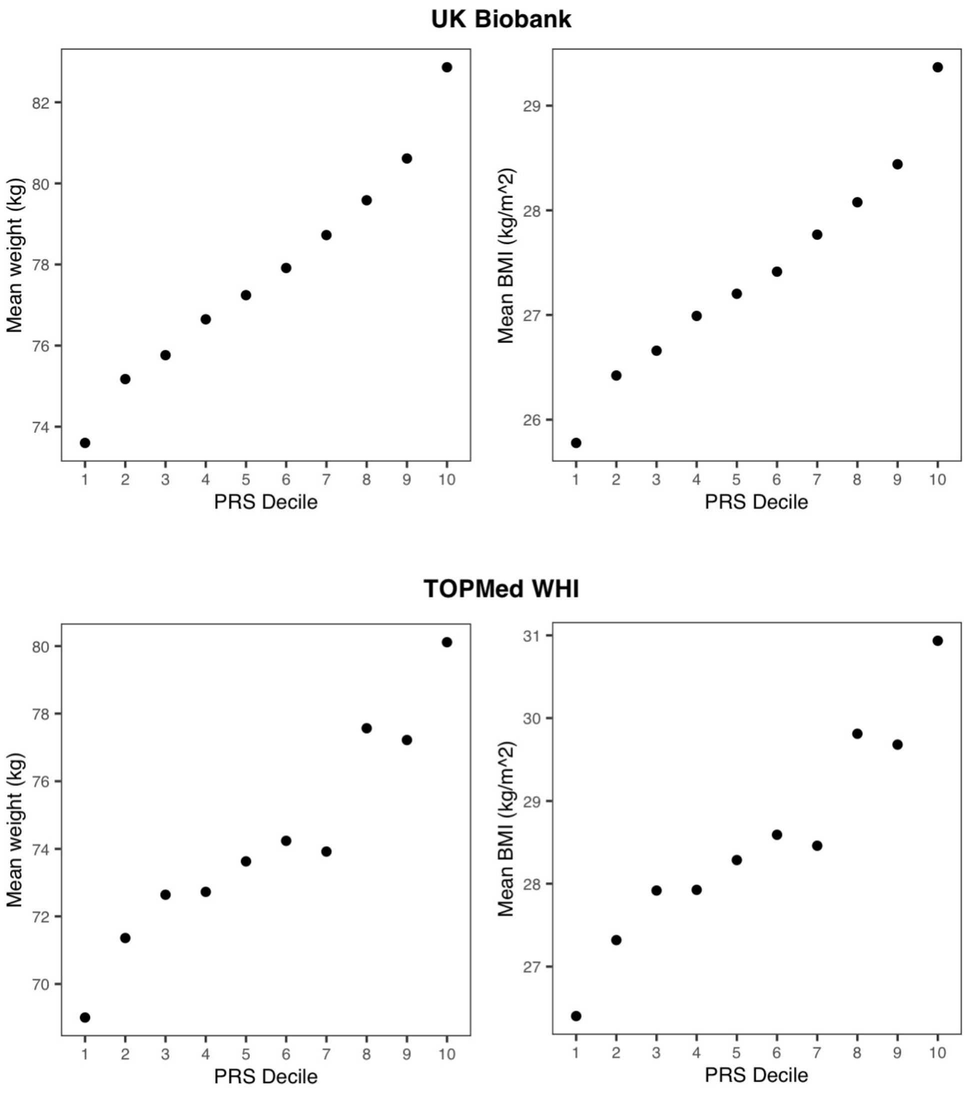


**Supplementary Figure 3.** Mean weight (left plots) and BMI (right plots) for individuals in the UKB validation cohort (top) and TOPMed WHI cohort (bottom), across deciles of a publicly-available polygenic risk score for BMI.

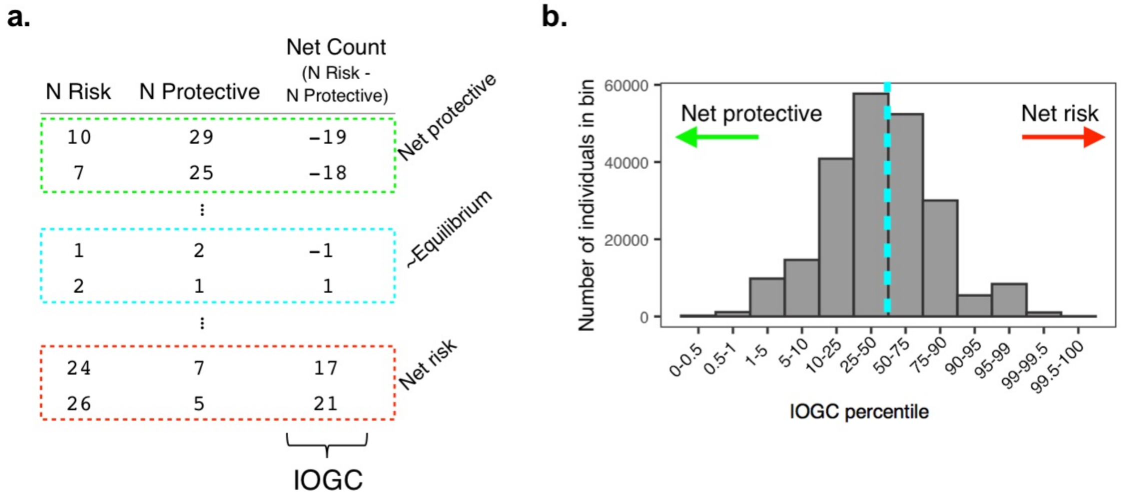


**Supplementary Figure 4.** Calculating outlier rare variant burden. **A.** Independent outlier gene count (IOGC) is defined as total outlier variants with a GWAS protective effect subtracted from total outlier variants with a GWAS risk effect, where outlier variants are collapsed to gene-level for individuals with >1 outlier variant per gene; **B.** Number of individuals in UKB across different percentiles of IOGC - individuals in bins to left of blue line have net-protective IOGC scores, whereas individuals in bins to the right have net-risk IOGC score.

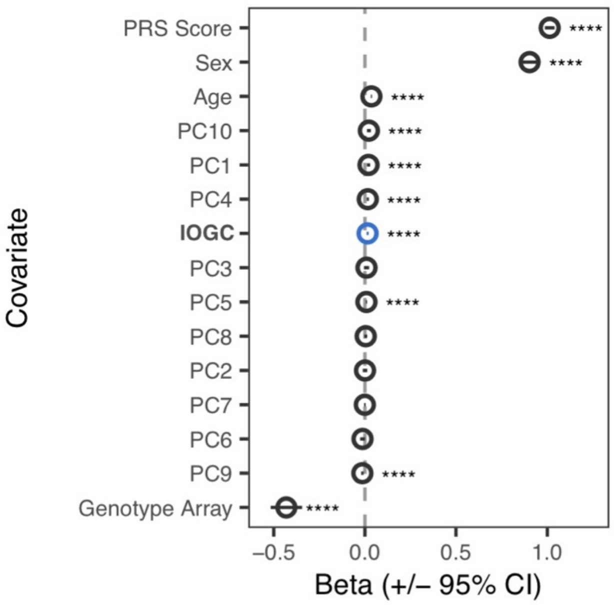


**Supplementary Figure 5.** Beta coefficients from a linear regression model testing the effect of IOGC score on body mass index, controlling for the effects of PRS, sex, age, genotype array, and the first 10 principal components of ancestry.

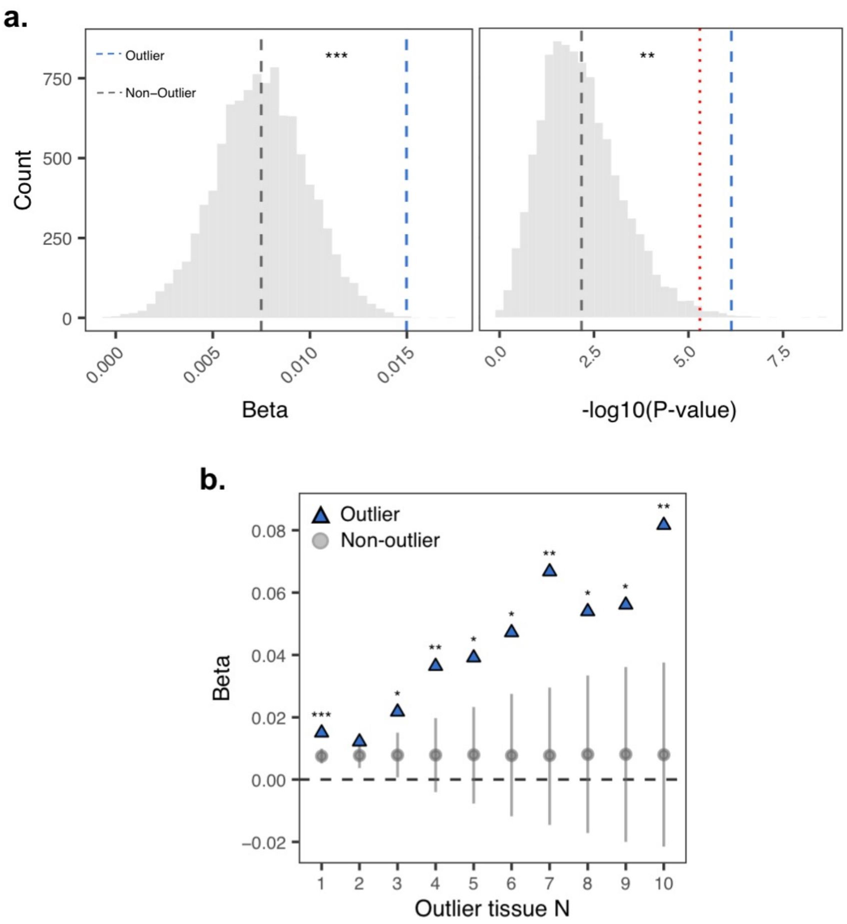


**Supplementary Figure 6. A.** Permutation test (N permutations = 10,000) comparing beta coefficients (left) and -log10(P-value) (right) for IOGC for outlier variants ("top-outlier" method, N variants = 8,272) (blue dashed line) and random samples of non-outlier variants (gray shading; gray dashed line indicates mean across permutations). Coefficients were estimated in a linear regression model controlling for the effects of PRS, sex, age, genotype array, and first 10 principal components of genetic ancestry. P-value is the empirical P-value. Red dotted line indicates the Bonferroni significance threshold. **B.** Permutation test as in **(A.)** as a function of multi-tissue outlier count. Coefficients for outlier variants are indicated in blue. Coefficients for non-outlier variants are summarized across permutations as mean (+/- SD). Non-outlier variants were randomly sampled to match the total number of outlier variants at each outlier tissue count threshold. P-values are empirical P-values.

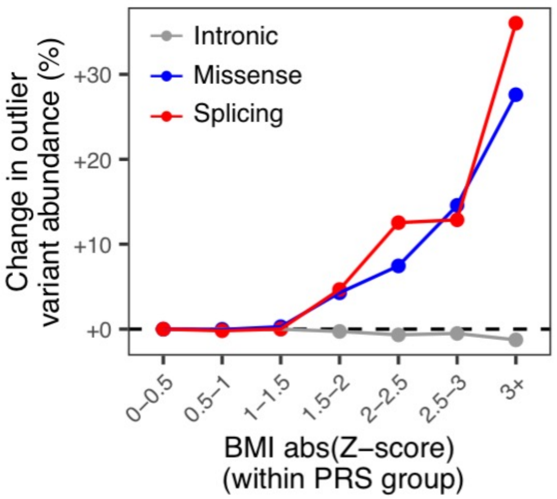


**Supplemental Figure 7.** Enrichment for missense and splicing outlier-associated variants in individuals with extreme IOGC score (bottom and top 10% of IOGC distribution), as a function of deviation from mean body mass index (calculated within each PRS group). Intron variants are shown as a control.

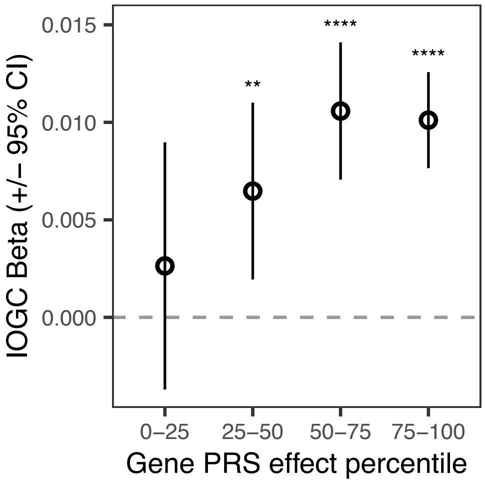


**Supplemental Figure 8.** IOGC score across percentiles of genes stratified by PRS effect weight. Each gene is summarized by the maximum effect size across all variants located within the gene. Higher percentiles comprise genes with larger PRS effects. IOGC score is increased in genes with larger effects on BMI.

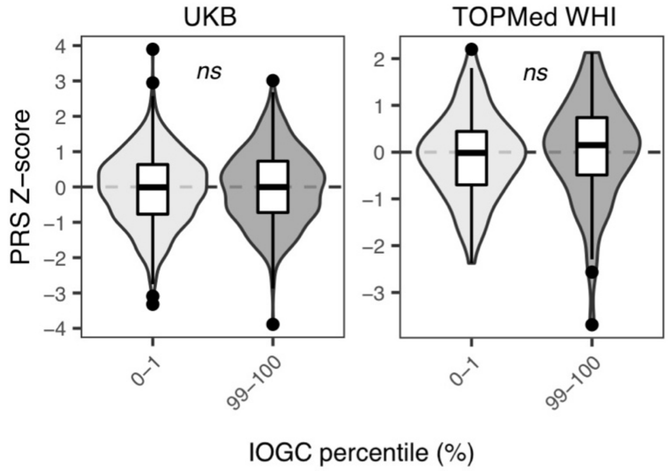


**Supplementary Figure 9.** Polygenic risk scores for UKB (left) and TOPMed WHI (right) individuals in top percentiles of IOGC score. No significant differences were observed in either cohort.

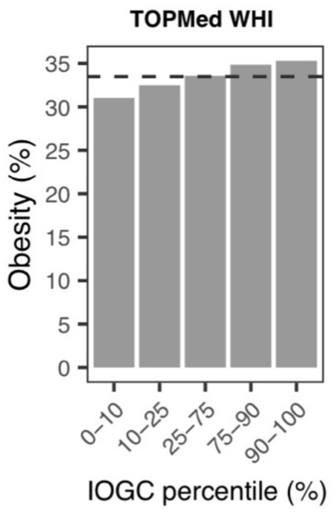


**Supplemental Figure 10.** Rates of obesity (BMI >= 30 kg/m$^2$) across TOPMed WHI individuals binned by percentiles of IOGC. Black dashed line indicates the mean rate of obesity in the cohort overall.

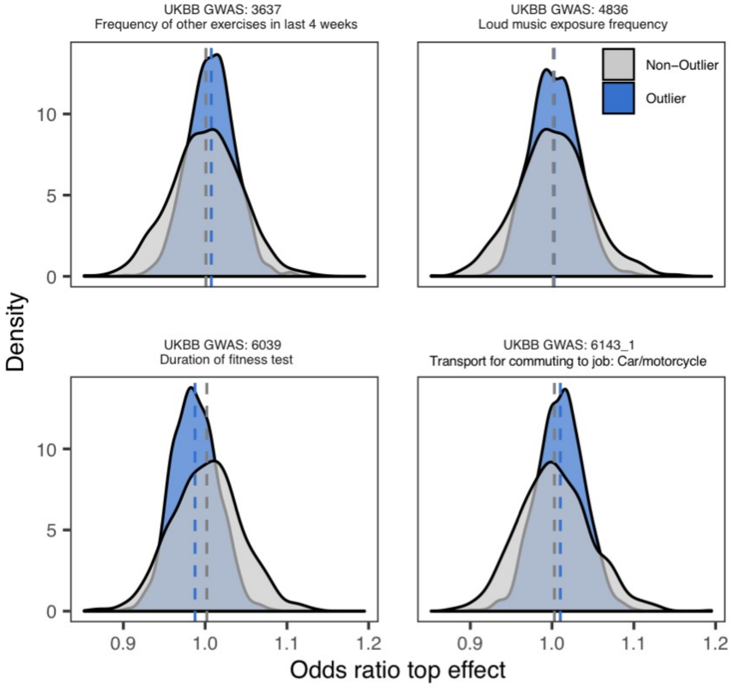


**Supplemental Figure 11.** Distribution of odds ratios from permutation testing (N permutations = 1,000) comparing GWAS absolute effect sizes of outlier vs. non-outlier variants (blue) and non-outlier variants (gray) across GWAS not expected to be as sensitive to gene outlier effects. Both distributions center around an odds ratio = 1, indicating no difference in GWAS absolute effect sizes of outlier and non-outlier variants.
